# Genome-wide investigation of rare germline copy-number variants in retinoblastoma

**DOI:** 10.1101/2024.11.19.24316720

**Authors:** Lesley M. Chapman Hannah, Jung Kim, Jazmyn L. Bess, Sungduk Kim, Paul S. Albert, Nathalie Japkowicz, Douglas R. Stewart, Zois Boukouvalas

## Abstract

Approximately 8,000 children are diagnosed with retinoblastoma (Rb) globally each year, and the rate of survival as well as prognosis can differ greatly based on access to quality screening and treatment. Over 90% of patients with the inherited bilateral form of Rb have germline variants *RB1*, whereas approximately 20-30% of the unilateral form of Rb harbor germline variants in *RB1*. In the following study, rare germline copy-number variants (CNVs) within and outside of the *RB1* gene were evaluated. Germline whole-genome sequencing (WGS) data from 134 Rb samples and 313 non-cancer controls of European ancestry were analyzed from the St. Jude Cloud. In an analysis of 1514 rare germline CNVs, non-negative matrix factorization (NMF) and Bayesian logistic regression identified 18 CNVs associated with Rb status. NMF analysis was used to reduce the high-dimensional feature space and resulted in 412 rare germline CNVs, one of which was found in *RB1*. A rare intronic germline CNV within the *ACDY9* gene (OR= 3.29, 95% CI = 0.56 to 6.63) as well as an event within the intronic region of the *PLXNC1* gene (OR= 2.24, 95% CI = 0.87 to 3.67) were found. In an evaluation of gene function within the UCSC hg38 Fetal Gene Atlas, *ACDY9* has a role in eye photoreceptor cell development, and *PLXNC1* has a role in eye horizontal cell development; both cell types have a functional role in Rb development. These findings suggest novel rare germline CNVs outside of the *RB1* gene could be associated with Rb risk.

## Introduction

Retinoblastoma (Rb) is a rare form of eye cancer that affects infants and young children^1,2^. Approximately 40% of Rb patients have the hereditary form of Rb. These individuals often present with the bilateral form of Rb that can be linked to germline variants in the *RB1* gene^2^ (OMIM: 180200), and are often at higher risk of subsequent malignant neoplasms^3^, and the remaining 60% have the unilateral form of Rb^2^. Greater than 80% of children with bilateral Rb have germline risk variants within the RB1 gene, while only 15% of unilateral Rb can be linked to germline variants in *RB1*^2,4^. Improved germline genetic screening could have a critical role in the early detection of Rb, which could improve disease surveillance and ultimately prognosis for many children susceptible to developing Rb. It has been shown that risk loci discovered via genome-wide association studies (GWAS) are limited in completely accounting for disease risk, and there is growing evidence that rare variants could have stronger effects on cancer risk compared to common variants^5,6^. Investigating variants within and outside of the *RB1* gene as well as diverse classes of variants are needed to better understand genes involved in Rb genetic risk. We evaluated the hypothesis that rare germline CNVs in genes outside of *RB1* that occur at a higher frequency in Rb cases versus cancer-free controls could have a role in contributing to Rb risk.

Copy-number variants (CNVs) are large-scale chromosomal aberrations that occur as a result of a duplication (amplification) or deletion of a segment of DNA^7^. CNVs can encompass 5-12% of the human genome^8–10^, and advances in high-throughput sequencing technologies have led to increased detection of these events. Several groups have identified germline CNVs as contributing to cancer predisposition. Deletions in *BRCA1* and *CHEK2* have been associated with breast cancer risk^11^, and several groups have identified germline CNVs within *RB1* associated with Rb risk^12^. A growing number of studies are identifying risk variants within and outside of *RB1*, but these searches have been limited to somatic changes^13^ or CNVs discovered within targeted gene panels and whole-exome data. In this study, we sought to identify recurrent rare germline CNVs in Rb patients within whole genome sequencing data from the St. Jude Cloud database. We implement variant discovery and filtering strategies as well as a dimensionality reduction approach to discover candidate CNVs associated with Rb risk. Non-negative matrix factorization (NMF) is a dimensionality reduction technique that reduces a high-dimensional feature space into two low-rank non-negative matrices - basis matrix (W) and the coefficient matrix (H)^14^. NMF incorporates non-negativity constraints to generate the lower ranking non-negative matrices, making the features within the matrices more interpretable^15,16^. The method has been used for various tasks such as: somatic signature discovery^17^, germline genomic analysis^18^, and feature selection^19,20^. A Bayesian logistic regression approach was chosen in order to most effectively analyze a small dataset as well as a dataset where the features (CNVs) are greater than the number of observations (cases and controls)^21,22^. In our current study, we use NMF to reduce the high-dimensional features space and Bayesian logistic regression to estimate the CNVs associated with Rb. Within this limited group of Rb patients and cancer-free controls, we were able to examine the relationship between rare germline CNVs and Rb, and to provide preliminary evidence for a set of novel germline CNVs that could contribute to an individual’s predisposition to Rb.

## Methods

### Study participants

Whole-genome sequencing (WGS) data from 134 individuals with retinoblastoma, and 313 cancer-free controls was collected from studies within the St. Jude Cloud initiative ^23^. Sample ancestry was defined using peddy (v. 0.4.8)^24^, and cases and controls were matched based on a weighted Mahalanobis distance metric using PCAmatchR (v 0.3.3)^25^. Matched case and control samples that were >80% European genetic ancestry were included for analysis (**Supplementary Figure 1**).

### Germline variant discovery and annotation

Germline copy number variants were detected within each individual sample Binary Alignment Map (BAM) file using three different variant callers: Manta (v1.6.0)^26^, Delly (v1.1.6)^27^, and CNVnator (v0.4.1)^28^, and SVtyper was used for joint genotyping. CNVs supported by all three callers were included for analysis. All variant calls were passing deletion calls with quality score greater than 30, and variants with minor allele frequency (MAF) <1% as determined by gnomAD (v4.0) were included for analysis. Quantile-quantile plots were generated to assess inflation (**Supplementary Figure 2**). All filtered CNVs were manually inspected using the Integrative Genomics Viewer (IGV) (v2.12.3)^29^, and CNVkit (v0.9.9)^30^ scatter plots and heatmaps. SURVIVOR^31^ was used to merge across all sample VCFs resulting in a filtered set of rare germline CNV deletions. AnnotSV (v3.4.2)^32^ was used to annotate the merged VCF. The approach for CNV discovery is summarized in **Figure 1**.

**Figure 1.**
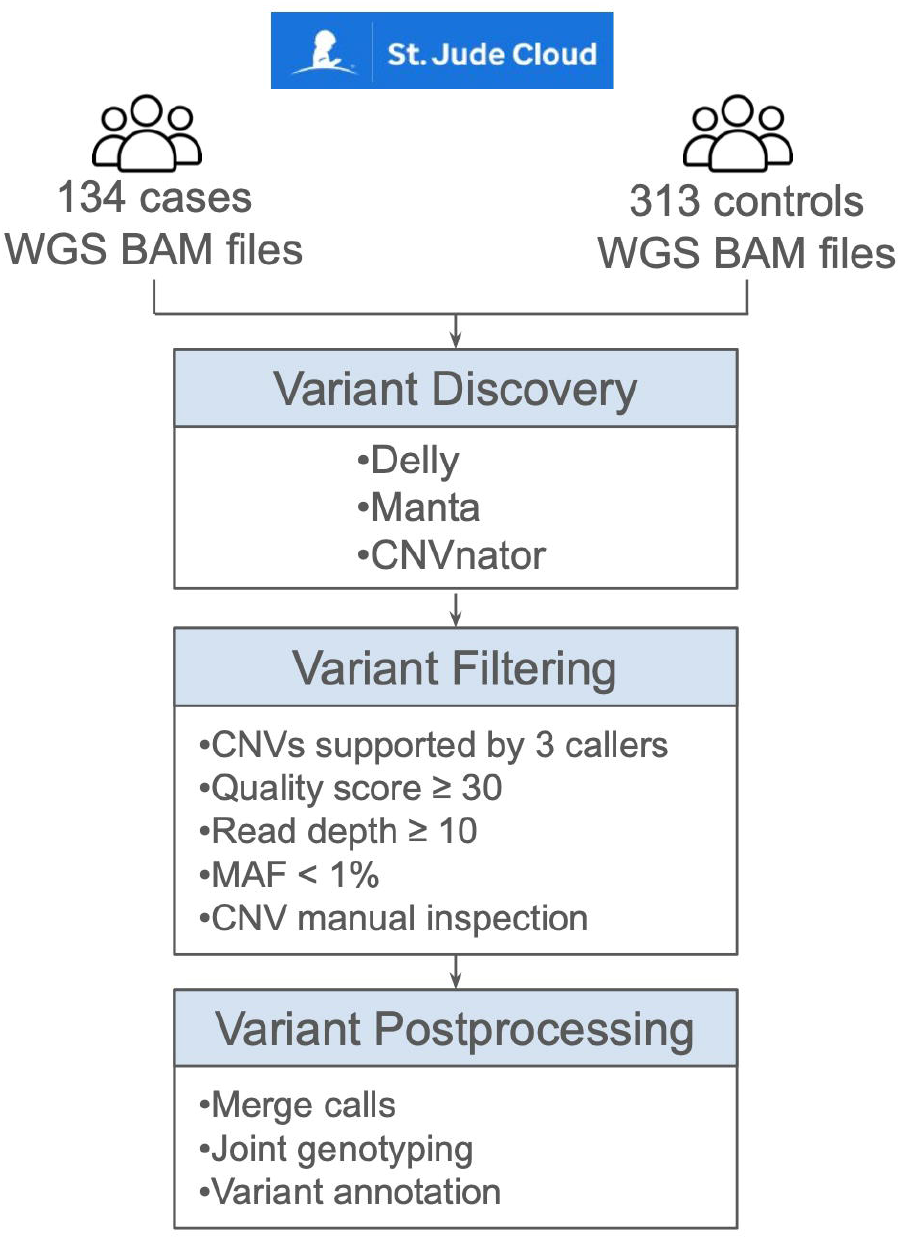
Schematic overview of the CNV discovery pipeline. Whole genome sequencing (WGS) barn files acquired from St. Jude Cloud Studies: Pediatric Cancer Genome Project (PCGP) and St. Jude LIFE (SJLIFE) for 134 retinoblastoma (Rb) cases and 313 cancer were analyzed. Three structural variant callers: Delly, Manta, CNVnator were applied for variant discovery. Quality-control filtering was used to remove low-confidence CNV calls. This resulted in 1514 rare germline CNVs for the 134 Rb cases and 313 controls.

### Statistical Analyses

Filtered CNV deletions were used as input for non-negative matrix factorization (NMF)^33^. NMF was applied to the filtered input data set [1514 CNV deletions and a total of 447 cases and controls] generating lower ranking matrices: basis matrix (W) and coefficient matrix (H). NMF was applied to the input data set using the Rcpp Machine Learning Library (RcppML) R library^16^. The CNVs with NMF weights greater than zero within the coefficient matrix were used as features to train the classifier. We fit Bayesian logistic regression with normal priors (mean 0, variance 1) using Rstan (v2.32.6) in order to infer odds ratios and 95% credible intervals (CI) for each variant. CNV deletions that do not contain the null effect of zero are considered to be associated with either the case or control label.

### Pathway analysis

All genes containing CNV deletions with NMF weights greater than zero were included for pathway analysis. Pathway analysis was conducted using PANTHER [Protein ANalysis THrough Evolutionary Relationships] (PANTHER version 19.0 Released 2024-06-19)^34^ and g:Profiler^35^, respectively. For each candidate gene list, PANTHER conducts a binomial test with multiple test correction (Bonferroni correction) that compares the candidate gene list to a reference list of genes to determine whether a set of Gene Ontology [GO] biological processes^36^. The web-based tool identifies pathways that contain significantly enriched genes based on Gene Ontology. For g:Profiler, p-values of the enriched pathways were determined using Fisher’s exact test with multiple hypothesis correction^37^. g:Profiler annotates gene groups based on the following GO categories: molecular functions (MF), cellular components (CC), and biological processes (BP)^35^.

## Results

### Overview of study cohort and rare germline copy number variant (CNV) discovery

The cohort consisted of 134 individuals with Rb and 313 cancer-free controls. Principal component analysis (PCA) analysis of the population structure identified a case-control set matched based on ancestry, and were all of European descent. For CNV detection, we performed genome-wide germline CNV discovery, genotyping, annotation and filtering jointly across all cases and controls. developed an approach that involved merging calls across three structural variant callers, variant merging, joint calling, and filtering. Our approach for CNV discovery is summarized in **Figure 1**. After sample- and variant-level quality control filtering, 1,514 rare germline CNV deletions were discovered with MAF < 1%. The average size of the rare germline CNV deletions was 6,902 base pairs (bp) with a size range of 51-253,990 bp (**Supplementary Figure 3**).

### Rare germline CNVs in Rb cohort and pathway analysis

Filtered CNVs were used as input for NMF analysis, and 412 rare germline CNVs were derived from the NMF coefficient matrix with weights greater than zero (**Supplementary Table 1**). There were 389 unique gene regions that harbored the 412 NMF-ranked CNVs, which were then evaluated for pathways that might be altered as a result of the presence of each CNVs. Genes with CNVs were evaluated using PANTHER and g:Profiler. g:Profiler pathway enrichment analysis of ranked genes identified 44 enriched GO biological processes including: nucleotide catabolic process (p-adj = 1.435×10^−2^), which has been shown to have a key role in regulating tumor development^38,39^, and cell development (p-adj = 3.267×10^−4^) (**Supplementary Table 2**). Upon further investigation of molecular function gene ontologies, beta-catenin binding (p-adj = 3.988×10^−3^) and cadherin binding (p-adj = 2.819×10^−2^) were also found to be significantly ranked (**Figure 2A**). The PANTHER pathway analysis determined that there were 15 CNV genes that are a part of the Wnt signaling pathway (p-value = 2.21×10^−4^, FDR = 3.53×10^−2^) (**Table 1, Supplementary Figure 4**). We then evaluated the relative abundance of the 15 CNVs involved in the Wnt signaling pathway within Rb cases and controls, and determined that Rb cases had a higher frequency of CNVs in Wnt signaling pathway genes compared to non-cancer controls (**Figure 2B**). The rare germline CNV counts were not statistically significant between cases and controls (**Figure 2B**).

**Figure 2.**
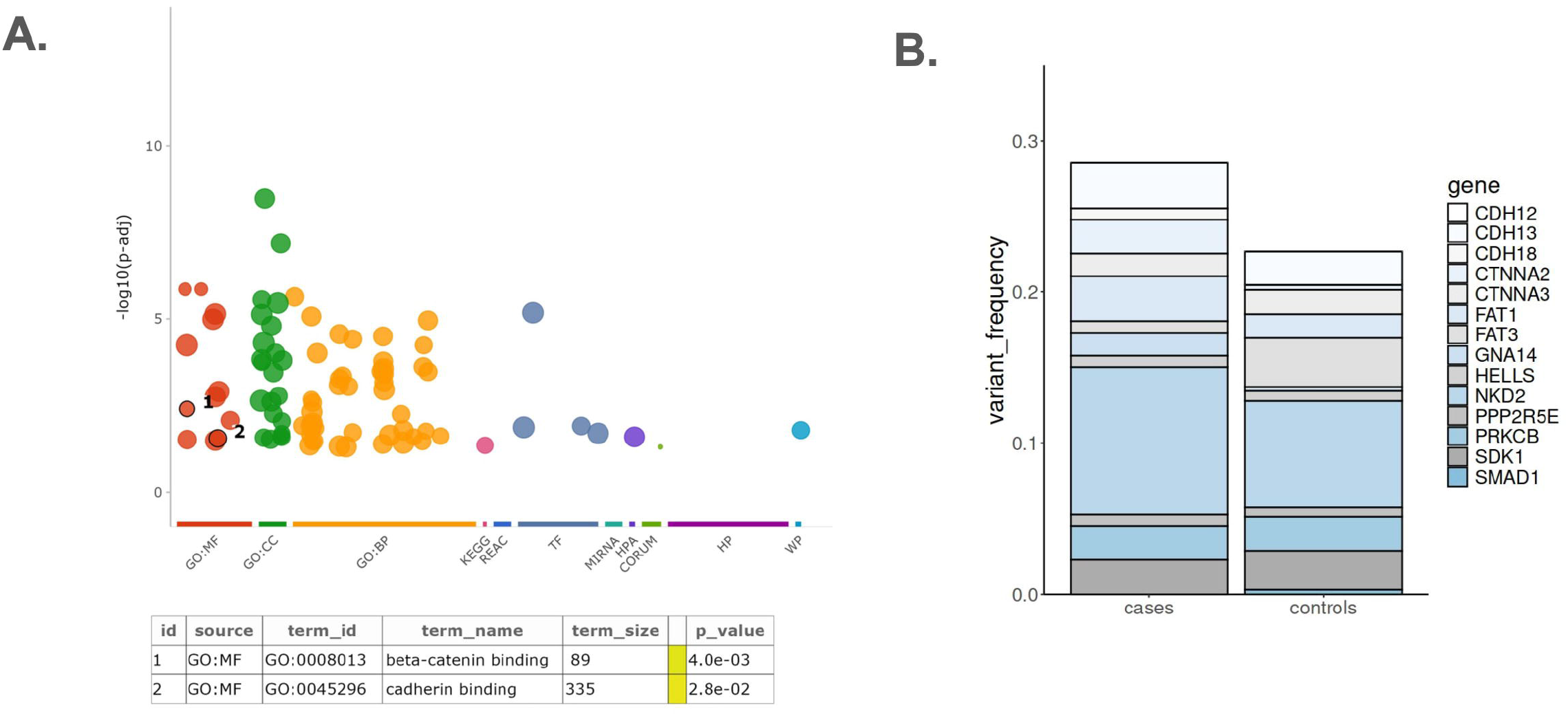
Functional enrichment analysis of genes with CNV deletions selected by NMF. A) g:Profiler Manhattan plot for genes harboring NMF selected CNV deletions. Lower panel shows gene enrichment analysis results for two gene ontology molecular function terms: beta-catenin binding and cadherin binding. B) Frequency of rare germline CNVs in Wnt signaling pathway genes in Rb cases and cancer-free controls.

**Table 1.** Pathway analysis of genes with CNV deletions selected by NMF. Pathway analysis for genes containing rare germline CNV deletions selected by NMF (n=389 genes) was completed using the PANTHER Classification System. One significant pathway was found: Wnt signaling pathway[PANTHER accession code P00057] (p-value(FDR corrected)= 3.53 × 10-^2^).

| PANTHER<br>Pathway(s) | Homo sapiens<br>pathway genes (n) | Genes<br>Observed (n) | Expected | Overrepresented(+)<br>Underrepresented(-) | Fold<br>Enrichment | Raw<br>p-value | FDR |
| --- | --- | --- | --- | --- | --- | --- | --- |
| Wnt signaling pathway | 306 | 15 | 5.06 | + | 2.97 | 2.21E-04 | 3.53E-02 |

**Table 2.** Rare germline CNVs associated with retinoblastoma(Rb) cases or cancer-free controls. Bayesian logistic regression applied to 1514 filtered rare germline CNV deletions in 134 Rb cases and 313 cancer-free controls. Parameter estimates include: mean effect (posterior mean) where posterior means higher than O represent variants associated with an Rb label and 95o/o credible intervals. Genes containing CNV deletions, CNV chromosome, start and end positions are reported.

| Gene | Chr | Start<br>(Build38) | End | Effect Size<br>(mean) | Lower CI<br>(2.5%) | Upper CI<br>(97.5%) |
| --- | --- | --- | --- | --- | --- | --- |
| AGBL4 | 1 | 49448841 | 49532907 | 1.95 | 0.19 | 3.84 |
| CLCA4 | 1 | 86562528 | 86573604 | -2.90 | -6.26 | -0.33 |
| KMO | 1 | 241569264 | 241580955 | 3.07 | 0.94 | 5.32 |
| IL1R2 | 2 | 102002681 | 102003129 | 4.44 | 1.71 | 7.56 |
| CAP2 | 6 | 17408955 | 17416154 | -1.08 | -1.87 | -0.35 |
| EXOC2 | 6 | 675519 | 676596 | -2.69 | -6.06 | -0.05 |
| AVL9 | 7 | 32510313 | 32511248 | -0.86 | -1.64 | -0.12 |
| SRRM3 | 7 | 76265824 | 76266169 | -1.02 | -1.78 | -0.30 |
| PIP | 7 | 143127749 | 143196815 | -3.15 | -6.54 | -0.44 |
| PTPRN2 | 7 | 158275950 | 158276407 | 0.64 | 0.08 | 1.21 |
| LOC100128993 | 8 | 19233998 | 19234404 | -1.64 | -2.64 | -0.73 |
| PLXNC1 | 12 | 94165580 | 94166314 | 2.24 | 0.87 | 3.68 |
| LOC100507065 | 12 | 65575974 | 65584706 | -0.70 | -1.36 | -0.06 |
| ADCY9 | 16 | 4084252 | 4091138 | 3.29 | 0.56 | 6.63 |
| NEDD4L | 18 | 58263874 | 58269908 | 1.50 | 0.20 | 2.79 |
| NMRK2 | 19 | 3939254 | 3940009 | 2.29 | 0.26 | 4.43 |
| SAMHD1 | 20 | 36894399 | 36896141 | 1.85 | 0.15 | 3.61 |
| LINC01692 | 21 | 24972164 | 24973450 | 2.85 | 0.51 | 5.43 |

### Rare germline CNV abundance in Rb cases and cancer-free controls

In order to estimate the association between the 412 NMF-ranked rare germline CNVs and Rb case-control status, Bayesian logistic regression was used to identify candidate CNVs. Our results identified 18 CNVs that had 95% credible intervals that did not contain the null effect (posterior mean of zero) (**Table 2**). Of these, a rare germline CNV in the intronic region of *ADCY9* gene (chr16:4084252-4091138) with a posterior mean of 3.29 (95% CI: [0.56,6.62]) was found in 3 Rb cases and 0 cancer-free controls (**Supplementary Figures 5-7**). Our analysis also identified an intronic CNV deletion in the *KMO* gene with a posterior mean of 3.07 (95% CI: [0.94,5.32]), and was found in 4 Rb cases and 2 non-cancer controls. Lastly, there were 3 Rb cases and 0 cancer-free controls that had a rare germline intronic CNV deletion in *IL1R2* (posterior mean= 4.44, 95% CI: [1.71-7.55]).

### Evaluation of intronic CNVs

We next evaluated select rare germline CNVs that occurred at a higher frequency in Rb cases to understand the role of intronic CNV deletions within somatic data from cBioPortal and the UCSC Browser. First, we assessed somatic CNVs in the *ADCY9* gene within the cBioPortal TCGA cohort. **Figure 3A** shows the distribution of individuals within TCGA with *ADCY9* CNV deletions and amplifications compared to samples with unaltered *ADCY9*. For somatic CNV deletions, it was found that 29 individuals had homozygous CNV deletions and 1701 individuals had heterozygous CNV deletions (**Figure 3A**). Our search also identified a 63 year old male with low grade glioma of European ancestry within the cBioPortal TCGA cohort that has a 21,247bp intronic CNV deletion in *ADCY9* [chr16:4,068,070-4,089,317] with a mean copy number (CN) log2 value of -0.746 and a tumor mRNA z-score of -0.746 (**Figure 3B**). To investigate the functional role of *ADCY9* in fetal development, we queried the *ADCY9* gene in the UCSC Browser hg38 Human Fetal Gene Atlas. The Human Fetal Gene Atlas consists of RNA-seq data from over 4-million single cells from 15 organs obtained during midgestational human development^47^. Our search determined that the *ADCY9* gene maximum mean gene expression value per cell (UMI/cell) was highest in photoreceptor cells compared to any other tissue type (0.62 UMI/cell) (**Figure 3C**). We also found a 734 bp rare germline CNV deletion in *PLXNC1* [chr12:94165580-94166314] that occurred at a higher frequency in Rb cases (posterior mean = 2.24, 95% CI: [0.87-3.67]). We then investigated somatic CNVs for *PLXNC1* within the same databases. The results showed that 8 individuals within the cBioPortal TCGA cohort had somatic deep CNV deletions and 1403 individuals had heterozygous (shallow) deletions in the *PLXNC1* gene (**Figure 4A**). Of the individuals with *PLXNC1* somatic deep CNV deletions, we identified a 74 year old female with glioblastoma who had a 4,452 bp intronic CNV deep deletion [chr12:94,549,550-94,554,002] in *PLXNC1*, which had a mean CN log2 value of -3.722 and no reported mRNA data (**Figure 4B**). We also found that the highest mean UMI/cell within fetal eye cells for *PLXNC1* was 0.77 expressed in eye horizontal cells (**Figure 4C**).

**Figure 3.**
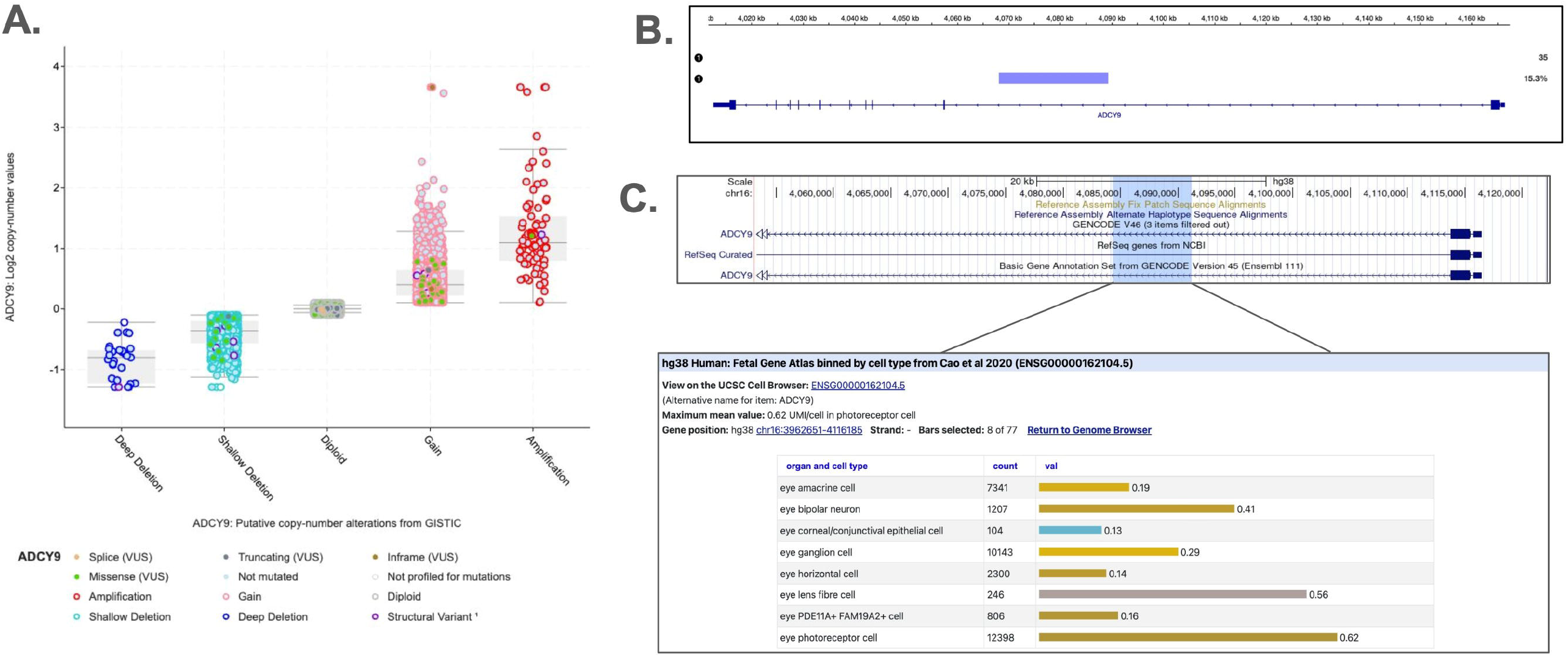
Evaluation of *ADCY9* copy number variants (CNVs) within cBioportal and the UCSC Browser. A) Frequency of CNV calls within the *ADCY9* gene in TCGA from cBioPortal. Each group represents GISTIC copy-number levels of each event within the *ADCY9* gene: Deep Deletion (homozygous deletion [n=29], Shallow Deletion (heterozygous deletion) [n=1701], Diploid (homozygous reference/no CNV event), Gain (heterozygous NV amplification) [n=2034], Amplification (homozygous CNV amplification)[n=82]. B) lntronic somatic deep deletion (chr16:4,084,252-4,091,138) [mean CN log2 value = -0.7459] found in a female TCGA patient with Glioblastoma Multiforme. C) Summary of hg38 Human Fetal Gene Atlas RNA abundance scores within fetal eye tissue derived from the UCSC Browser.

**Figure 4.**
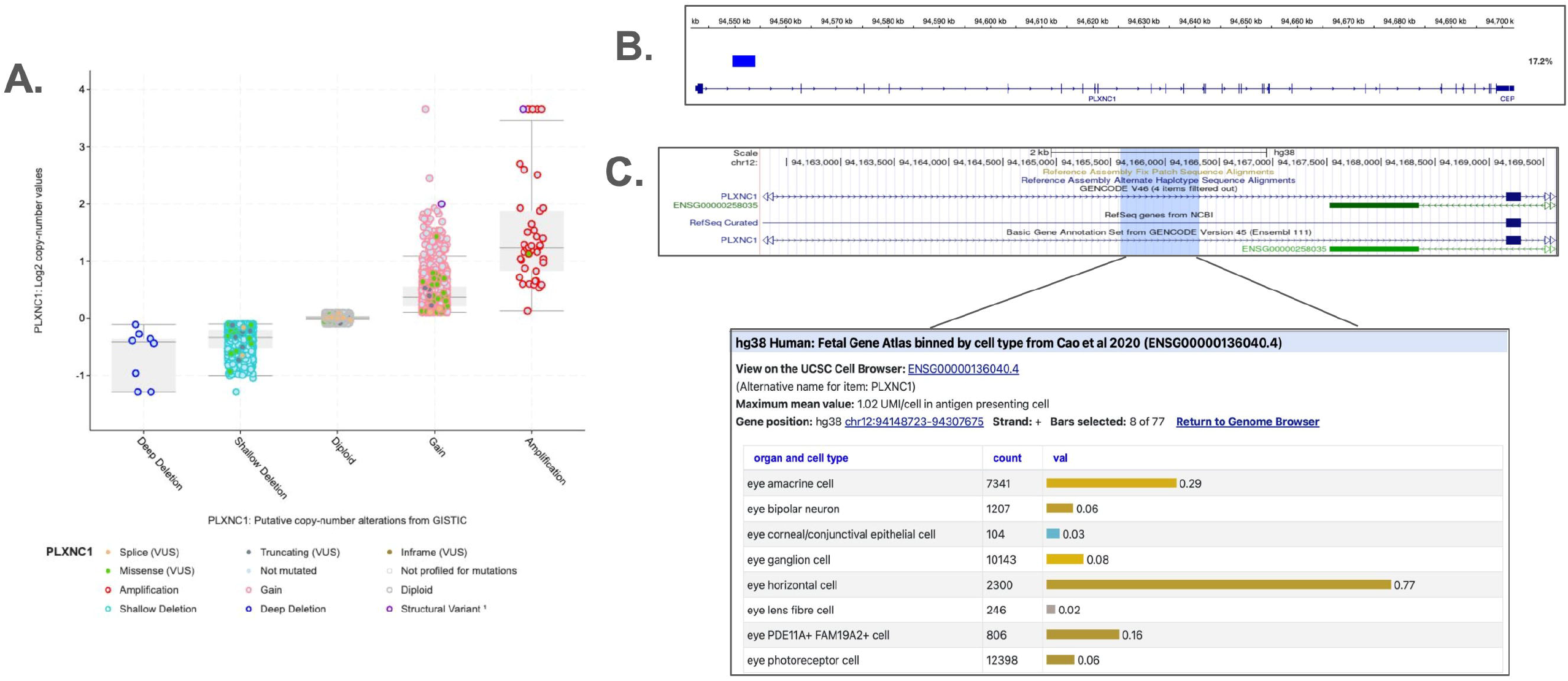
Evaluation of *PLXNC1* copy number variants (CNVs) within cBioportal and the UCSC Browser. A) Frequency of CNV calls within the *PLXNC1* gene in TCGA from cBioPortal. Each group represents GISTIC copy-number levels of each event within the *PLXNC1* gene: Deep Deletion (homozy ous deletion) [n=B], Shallow Deletion (heterozygous deletion) [n=1403], Diploid (homoz gous reference/no CNV event, Gain (heterozygous CNV amplification) [n=1766], Amplification (homozygous CNV ampli ication)[n=42]. B) lntronic somatic deep deletion (chr12:94,549,550-94,554,002) [mean CN log2-value = -3.7224] found in a female TCGA patient with Glioblastoma Multiforme. C) Summary of hg38 Human Fetal Gene Atlas KNA abundance scores within fetal eye tissue derived from the UCSC Browser.

## Discussion

In this study, we surveyed rare germline CNV deletions across the whole genome within a set of Rb cases and cancer-free controls from the St. Jude Cloud database. Here we describe rare germline CNVs outside of the *RB1* gene that could potentially contribute to retinoblastoma risk. Our initial NMF analysis of rare germline CNVs identified 412 ranked CNVs, and a pathway analysis of genes harboring these variants identified 16 genes within the Wnt signaling pathway. Rare germline CNVs within Wnt signaling pathway genes occurred at a higher frequency in Rb cases versus cancer-free controls, although found to be not statistically significant between the two groups. Bayesian logistic regression analysis of the 412 NMF ranked CNVs identified 18 statistically significant rare germline CNVs that occurred at a higher frequency in Rb cases versus cancer-free controls.

The genes containing NMF ranked CNVs included a CNV found in the *RB1* gene as well as a CNV deletion found in the *RB1* Inducible Coiled-Coil 1 [*RB1CC1*] gene, both of which were only found in one individual with Rb, respectively, but were not found to be statistically significant. However, this provides some evidence that the NMF analysis had biologically relevant findings relevant to the Rb phenotype. Even though most candidate CNVs identified by Bayesian logistic occurred at a low frequency in Rb cases, CNVs found in select candidate genes could have a role in cancer development. Of these, the *ADCY9* gene has been shown to have a role in eye development as well as cancer progression. The adenylate cyclase (ADCY) gene family are genes that encode for a group of glycoproteins that regulate intracellular signaling, and have regulatory roles in a number of biological processes including: angiogenesis, apoptosis, and cell proliferation ^48^. *ADCY9* is an adenylyl cyclase that converts ATP to cyclic AMP^49^, and has been found to act as a tumor suppressor within lung cancer^50^. Other members of the ADCY gene family have been shown to have roles in regulating other cancers such as a CNV deletion found in breast cancer within *ADCY8* ^51^. Also, single cell RNAseq data from the Human Fetal Gene Atlas suggests that *ADCY9* expression has a key role in photoreceptor cell development, a cell type involved in the early stages of Rb initiation^52^. Variants within the *ADCY9* gene have also been linked to pediatric ocular diseases such as - retinopathy of prematurity - an ocular disease that can occur in premature infants^49^. This suggests that germline CNV mutations in the *ADCY9* gene could also contribute to Rb risk. CNVs within *PLXNC1* also occurred at a higher frequency in Rb cases, and has been shown to act as a tumor suppressor in melanoma^53^. This could also be an additional candidate gene associated with Rb risk. The results of this study were generated based on a cohort with a small sample size. Although this a limitation of the study, the results help provide evidence for the need for future studies with larger sample sizes to further validate the frequency of candidate risk CNVs within and outside of the *RB1* gene. Lastly, this study focused on the evaluation of rare germline CNV deletions associated with Rb risk, which is largely due to other classes of variants (i.e.: amplifications) being comparatively more difficult to discover because of limitations in short read WGS technologies as well as variant discovery tools^54^.

Future studies that utilize more advanced sequencing and variant discovery approaches are needed to discover additional classes of structural variants that could be associated with Rb risk. Also, many of the CNVs identified as a part of this analysis occur within intronic regions of genes, additional experiments are needed to further define the functional impact of these CNVs on gene function. Individuals of non-European ancestry often have a greater number of variants of unknown significance [VUS] that can be recategorized to better estimate disease risk^55^. Future studies that investigate rare germline risk in Rb cases on non-European ancestry are needed. Additional studies investigating rare germline variants that are underexplored in Rb are needed, such as single nucleotide variants within noncoding regions of the genome. These studies could help provide additional insight on Rb genetic predisposition.

## Data Availability

Whole genome sequencing BAM files and phenotype data used for this study are available upon request via St. Jude Cloud (https://www.stjude.cloud/).

## Acknowledgements

This study was supported by the Intramural Research Program of the Division of Cancer Epidemiology and Genetics, National Cancer Institute, and National Institutes of Health. This research utilized the high performance computing cluster resources of the National Institutes of Health (NIH, USA; Biowulf, http://hpc.nih.gov).

## Notes

### Competing Interest Statement

The authors have declared no competing interest.

### Author Declarations

All data used for this study were made available via St. Jude Cloud (https://www.stjude.cloud/), and were made publicly available before the initiation of this study.

## References

1. Lin, F. Y. & Chintagumpala, M. M. Neonatal Retinoblastoma. Clin. Perinatol. 48, 53–70 (2021).

2. Marković, L., Bukovac, A., Varošanec, A. M., Šlaus, N. & Pećina-Šlaus, N. Genetics in ophthalmology: molecular blueprints of retinoblastoma. Hum. Genomics 17, 82 (2023).

3. Fabius, A. W. M., van Hoefen Wijsard, M., van Leeuwen, F. E. & Moll, A. C. Subsequent Malignant Neoplasms in Retinoblastoma Survivors. Cancers 13, 1200 (2021).

4. Afshar, A. R. et al. Next-Generation Sequencing of Retinoblastoma Identifies Pathogenic Alterations beyond RB1 Inactivation That Correlate with Aggressive Histopathologic Features. Ophthalmology 127, 804–813 (2020).

5. Mitchell, J. et al. Characterising the contribution of rare protein-coding germline variants to prostate cancer risk and severity in 37,184 cases. medRxiv 2024.05.10.24307164 (2024) doi:10.1101/2024.05.10.24307164.

6. Claussnitzer, M. et al. A brief history of human disease genetics. Nature 577, 179–189 (2020).

7. Freeman, J. L. et al. Copy number variation: New insights in genome diversity. Genome Res. 16, 949–961 (2006).

8. Zhang, F., Gu, W., Hurles, M. E. & Lupski, J. R. Copy Number Variation in Human Health, Disease, and Evolution. Annu. Rev. Genomics Hum. Genet. 10, 451–481 (2009).

9. Copy Number Variation | Learn Science at Scitable. http://www.nature.com/scitable/topicpage/copy-number-variation-445.

10. McCarroll, S. A. et al. Integrated detection and population-genetic analysis of SNPs and copy number variation. Nat. Genet. 40, 1166–1174 (2008).

11. Easton, D. F. et al. Gene-Panel Sequencing and the Prediction of Breast-Cancer Risk. N. Engl. J. Med. 372, 2243–2257 (2015).

12. Lan, X. et al. Spectrum of RB1 Germline Mutations and Clinical Features in Unrelated Chinese Patients With Retinoblastoma. Front. Genet. 11, (2020).

13. Kooi, I. E. et al. Somatic genomic alterations in retinoblastoma beyond RB1 are rare and limited to copy number changes. Sci. Rep. 6, 25264 (2016).

14. Al-Ghafer, I. A. et al. NMF-guided feature selection and genetic algorithm-driven framework for tumor mutational burden classification in bladder cancer using multi-omics data. Netw. Model. Anal. Health Inform. Bioinforma. 13, 26 (2024).

15. Kim, H. & Park, H. Sparse non-negative matrix factorizations via alternating non-negativity-constrained least squares for microarray data analysis. Bioinformatics 23, 1495–1502 (2007).

16. DeBruine, Z. J., Pospisilik, J. A. & Triche, T. J. Fast and interpretable non-negative matrix factorization for atlas-scale single cell data. 2021.09.01.458620 Preprint at 10.1101/2021.09.01.458620 (2024).

17. Alexandrov, L. B. et al. Signatures of mutational processes in human cancer. Nature 500, 415–421 (2013).

18. Germline genomic patterns are associated with cancer risk, oncogenic pathways, and clinical outcomes | Science Advances. https://www.science.org/doi/10.1126/sciadv.aba4905.

19. Zeng, Z. et al. Cancer classification and pathway discovery using nonnegative matrix factorization. J. Biomed. Inform. 96, 103247 (2019).

20. Yu, N., Gao, Y.-L., Liu, J.-X., Wang, J. & Shang, J. Robust hypergraph regularized non-negative matrix factorization for sample clustering and feature selection in multi-view gene expression data. Hum. Genomics 13, 46 (2019).

21. van de Schoot, R., Broere, J. J., Perryck, K. H., Zondervan-Zwijnenburg, M. & van Loey, N. E. Analyzing small data sets using Bayesian estimation: the case of posttraumatic stress symptoms following mechanical ventilation in burn survivors. Eur. J. Psychotraumatology 6, 10.3402/ejpt.v6.25216 (2015).

22. Chakraborty, S., Ghosh, M. & Mallick, B. K. Bayesian nonlinear regression for large p<math><mi is=“true”>p</mi></math> small n<math><mi is=“true”>n</mi></math> problems. J. Multivar. Anal. 108, 28–40 (2012).

23. McLeod, C. et al. St. Jude Cloud: A Pediatric Cancer Genomic Data-Sharing Ecosystem. Cancer Discov. 11, 1082–1099 (2021).

24. Pedersen, B. S. & Quinlan, A. R. Who’s Who? Detecting and Resolving Sample Anomalies in Human DNA Sequencing Studies with Peddy. Am. J. Hum. Genet. 100, 406–413 (2017).

25. Brown, D. W., Myers, T. A. & Machiela, M. J. PCAmatchR: a flexible R package for optimal case–control matching using weighted principal components. Bioinformatics 37, 1178–1181 (2021).

26. Manta: rapid detection of structural variants and indels for germline and cancer sequencing applications | Bioinformatics | Oxford Academic. https://academic.oup.com/bioinformatics/article/32/8/1220/1743909.

27. Rausch, T. et al. DELLY: structural variant discovery by integrated paired-end and split-read analysis. Bioinformatics 28, i333–i339 (2012).

28. Abyzov, A., Urban, A. E., Snyder, M. & Gerstein, M. CNVnator: An approach to discover, genotype, and characterize typical and atypical CNVs from family and population genome sequencing. Genome Res. 21, 974–984 (2011).

29. Robinson, J. T. et al. Integrative genomics viewer. Nat. Biotechnol. 29, 24–26 (2011).

30. Talevich, E., Shain, A. H., Botton, T. & Bastian, B. C. CNVkit: Genome-Wide Copy Number Detection and Visualization from Targeted DNA Sequencing. PLOS Comput. Biol. 12, e1004873 (2016).

31. Jeffares, D. C. et al. Transient structural variations have strong effects on quantitative traits and reproductive isolation in fission yeast. Nat. Commun. 8, 14061 (2017).

32. Geoffroy, V. et al. AnnotSV: an integrated tool for structural variations annotation. Bioinformatics 34, 3572–3574 (2018).

33. Lee, D. D. & Seung, H. S. Learning the parts of objects by non-negative matrix factorization. Nature 401, 788–791 (1999).

34. Mi, H., Muruganujan, A. & Thomas, P. D. PANTHER in 2013: modeling the evolution of gene function, and other gene attributes, in the context of phylogenetic trees. Nucleic Acids Res. 41, D377–D386 (2013).

35. Reimand, J., Kull, M., Peterson, H., Hansen, J. & Vilo, J. g:Profiler—a web-based toolset for functional profiling of gene lists from large-scale experiments. Nucleic Acids Res. 35, W193–W200 (2007).

36. PANTHER Pathway: an ontology-based pathway database coupled with data analysis tools -PMC. https://www.ncbi.nlm.nih.gov/pmc/articles/PMC6608593/.

37. Reimand, J. et al. Pathway enrichment analysis and visualization of omics data using g:Profiler, GSEA, Cytoscape and EnrichmentMap. Nat. Protoc. 14, 482–517 (2019).

38. Ali, E. S. & Ben-Sahra, I. Regulation of nucleotide metabolism in cancers and immune disorders. Trends Cell Biol. 33, 950–966 (2023).

39. Wu, H. et al. Targeting nucleotide metabolism: a promising approach to enhance cancer immunotherapy. J. Hematol. Oncol.J Hematol Oncol 15, 45 (2022).

40. Lin, W.-H., Cooper, L. M. & Anastasiadis, P. Z. Cadherins and catenins in cancer: connecting cancer pathways and tumor microenvironment. Front. Cell Dev. Biol. 11, 1137013 (2023).

41. Tell, S., Yi, H., Jockovich, M.-E., Murray, T. G. & Hackam, A. S. The Wnt signaling pathway has tumor suppressor properties in retinoblastoma. Biochem. Biophys. Res. Commun. 349, 261–269 (2006).

42. Su, Y.-J., Chang, Y.-W., Lin, W.-H.Liang, C.-L. & Lee, J.-L. An aberrant nuclear localization of E-cadherin is a potent inhibitor of Wnt/β-catenin-elicited promotion of the cancer stem cell phenotype. Oncogenesis 4, e157–e157 (2015).

43. Herzig, M., Savarese, F., Novatchkova, M., Semb, H. & Christofori, G. Tumor progression induced by the loss of E-cadherin independent of β-catenin/Tcf-mediated Wnt signaling. Oncogene 26, 2290–2298 (2007).

44. Yu, W., Yang, L., Li, T. & Zhang, Y. Cadherin Signaling in Cancer: Its Functions and Role as a Therapeutic Target. Front. Oncol. 9, (2019).

45. Liu, J. et al. Wnt/β-catenin signalling: function, biological mechanisms, and therapeutic opportunities. Signal Transduct. Target. Ther. 7, 1–23 (2022).

46. Valenta, T., Hausmann, G. & Basler, K. The many faces and functions of β-catenin. EMBO J. 31, 2714–2736 (2012).

47. Cao, J. et al. A human cell atlas of fetal gene expression. Science 370, eaba7721 (2020).

48. Guo, R. et al. Targeting Adenylate Cyclase Family: New Concept of Targeted Cancer Therapy. Front. Oncol. 12, (2022).

49. Teixeira, H. M. P., Alcantara-Neves, N. M., Barreto, M., Figueiredo, C. A. & Costa, R. S. Adenylyl cyclase type 9 gene polymorphisms are associated with asthma and allergy in Brazilian children. Mol. Immunol. 82, 137–145 (2017).

50. Tang, Y. et al. ADCY9 functions as a novel cancer suppressor gene in lung adenocarcinoma. J. Thorac. Dis. 15, (2023).

51. Dennis, J. et al. Rare germline copy number variants (CNVs) and breast cancer risk. Commun. Biol. 5, 1–15 (2022).

52. Bremner, R. & Sage, J. The origin of human retinoblastoma. Nature 514, 312–313 (2014).

53. Lazova, R., Gould Rothberg, B. E., Rimm, D. & Scott, G. The Semaphorin 7A Receptor Plexin C1 Is Lost During Melanoma Metastasis. Am. J. Dermatopathol. 31, 177 (2009).

54. Mahmoud, M. et al. Structural variant calling: the long and the short of it. Genome Biol. 20, 246 (2019).

55. Tischler, J., Crew, K. D. & Chung, W. K. The role of tumor and germline genetic testing in breast cancer management. Ann. Intern. Med. 171, 925–930 (2019).

